# Antibody responses to known and unknown SARS-CoV-2 infections after mRNA vaccine booster

**DOI:** 10.1101/2022.05.06.22274719

**Authors:** Alexis R. Demonbreun, Amelia Sancilio, Lauren A. Vaught, Nina L. Reiser, Lorenzo Pesce, Eoin P. Sode, Brian Mustanski, Richard D’Aquila, Elizabeth M. McNally, Thomas W. McDade

## Abstract

We followed a fully-vaccinated (two mRNA vaccine doses) community cohort (n=41) without prior COVID-19 diagnosis from September 2021 through March 2022 through the Omicron wave following a booster mRNA vaccination. 19.5% of participants reported a known SARS-CoV-2 infection during the Omicron wave, which was confirmed by anti-nucleocapsid IgG. An additional 36.5% also developed anti-nucleocapsid IgG after the boost, consistent with unknown, asymptomatic SARS-CoV-2 infection during this period. Infection defined by anti-nucleocapsid IgG, whether known to participant or not, increased anti-spike IgG levels, relative to those lacking anti-nucleocapsid IgG, at 120 days post-booster.

## Introduction

It is well-established that COVID-19 vaccinations generate robust cellular- and antibody-mediated immune responses that provide lasting protection against severe disease [1, 2]. However, levels of neutralizing antibodies decline over time, and antigenic shifts in novel variants reduce the effectiveness of antibody-mediated neutralization, resulting in higher rates of breakthrough infections, the vast majority of which are mild [3-5]. This dynamic has played out dramatically with Omicron (B.1.1.529), the most recent variant to sweep over the US beginning in late 2021, peaking in January 2022 with the highest daily case rates recorded since the beginning of the pandemic [6]. The Centers for Disease Control and Prevention estimates that by the end of February 2022, 57.7% of the US Population had serological evidence of natural infection with SARS-CoV-2 [4].

In this study, we examine the antibody response to natural infection during the Omicron wave, as well as the antibody response to booster doses of mRNA vaccines. Before the arrival of Omicron we recruited a cohort of adults in Chicago with no history of clinically diagnosed COVID-19, who were fully vaccinated (two doses of Pfizer or Moderna vaccine), and who indicated the intent to receive a third booster dose. The follow up period of the study coincided with the arrival of the Omicron variant, which was first detected in the Chicagoland area December 7, 2021 and quickly became the dominant strain, with cases subsiding February 2022. With blood sampling prior to and one week after the administration of the booster dose, as well as follow up sampling for up to four months during the peak of Omicron, we document the magnitude of antibody response to the booster and model the rate of decline over time.

In addition, by measuring antibodies against the SARS-CoV-2 nucleocapsid protein, we are able to detect responses triggered by infection with the SARS-CoV-2 virus itself and not the vaccine. By measuring anti-nucleocapsid (N), we can detect past infection even if participants were asymptomatic and/or not identified as infected with clinical or at-home tests for COVID-19 [7]. We can therefore estimate the prevalence of infection after a booster dose during the Omicron wave and also compare the antibody response to boosting with and without subsequent infection. Despite large increases in anti-spike IgG concentrations recognizing wild-type, Delta, and Omicron variants of SARS-CoV-2 seven days after the booster, anti-spike antibody concentrations declined over the subsequent four months. Notably, 56% of participants developed anti-N antibodies indicating infection by the end of that follow up period. At 120 days after a boost, anti-spike IgG concentrations recognizing these variants were 2.5 to 4.7 times higher in those with serological evidence of infection after a boost than in those who did not develop anti-N antibody. These results suggest that Omicron infection after a booster vaccination may generate stronger or longer lasting antibody-mediated protection against future re-infection.

## Methods

All research activities were implemented under protocols approved by Northwestern University’s institutional review board (#STU00212457 and #STU00212472).

Participants were recruited from a previously conducted community-based serosurvey across the Chicagoland area, Screening for Coronavirus Antibodies in Neighborhoods (SCAN) [8]. Participants e-consented and provided information regarding COVID-19 viral history and vaccination status. All participants had received only 2 doses of Pfizer or Moderna mRNA vaccine (initial series) and reported intent to receive a third (booster) vaccine dose. None reported being immunocompromised or receiving a COVID-19 diagnosis prior to their third vaccine dose. Finger stick dried blood spot (DBS) samples were self-collected September 2021 through March 2022, immediately prior to booster administration (pre-sample), 6-10 days after receiving mRNA booster vaccine, and then at approximately 60, 90, and 120 days after the booster dose. Thirty-three participants provided a complete series of samples. An additional eight participants provided pre-booster and 120-day samples only.

Concentrations of anti-SARS-CoV-2 IgG antibodies against spike and nucleocapsid (N) antigens were quantified using the Meso Scale Discovery multiplex anti-IgG electrochemiluminescence assay (SARS-CoV-2 Panel 24, K15575U), which was specifically optimized and validated for use with DBS [9]. Anti-spike IgG antibodies against wild-type (WT; Wuhan A) SARS-CoV-2 were quantified, as well as IgG antibodies against Delta (B.1.617.2; AY.2) and Omicron variants (B.1.1.529; BA.1). Samples were classified as N-positive when the IgG concentration was greater than 3 standard deviations above the mean of pre-pandemic (negative samples), determined as 0.19 (AU/mL). If the pre-sample was N-positive, then a new infection was determined if the subsequent N IgG concentration increased compared to the pre-sample. Samples were analyzed in duplicate, with the average result reported.

Statistics were performed using a two-way repeated measures ANOVA with Tukey’s multiple comparison or Friedman test, with p<0.05 use as the criterion for statistical significance.

## Results

The sample included approximately equal numbers of women and men, with a median age of 41 years (**Supplemental Table 1**). The median duration between second mRNA vaccine dose and booster was 120 days. Participants were categorized into three groups based on anti-N IgG serological results and COVID-19 diagnosis history: known COVID-19 positive (self-reported COVID19+ diagnosis and serologically-confirmed infection by anti-N positivity at any point after pre-booster DBS sampling), unknown COVID-19 positive (lack of positive antigen or PCR test, serologically-confirmed infection by anti-N positivity at any point after pre-booster DBS sampling), and SARS-CoV-2 negative (lack of positive antigen or PCR test, serologically-confirmed anti-N negativity from all pre- and post-booster samples).

Immediately prior to booster vaccination (pre-sample), anti-N and anti-spike WT, Delta, and Omicron IgG levels were not significantly different across the three groups (**Figure 1**). Over the subsequent four months all participants received a booster dose, and the Omicron wave swept over Chicago. Eight of 41 participants (19.5%) reported a post-booster positive COVID-19 test, with serologically confirmed natural infection for all. No COVID-19+ participants required medical care. Fifteen of 41 participants (36.5%) showed serological evidence of natural infection after the booster dose, but did not report a positive COVID-19 test. Anti-N IgG levels increased significantly for both groups, with a larger increase in the known COVID-19+ group in comparison with the unknown COVID-19+ group.

**Figure 1.**
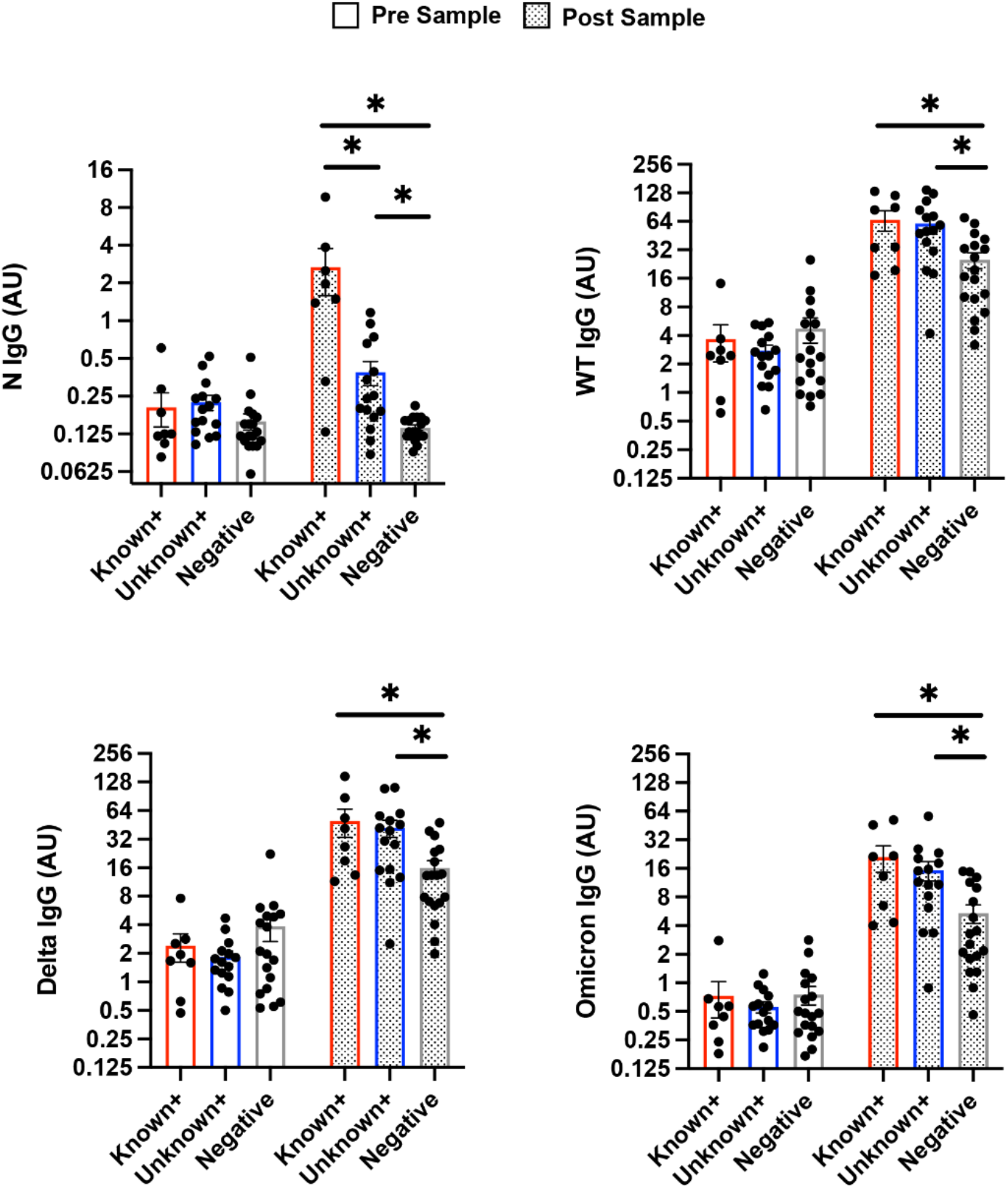
Anti-spike IgG antibody response to infection is similar between known COVID-19 positive and serologically-confirmed unknown COVID-19 positive groups. Plots show individual level data of matched samples before booster vaccination prior to the start of the Omicron wave (pre) and approximately 120 days post booster vaccination at the end of the Chicagoland Omicron Wave (post). Participants were categorized into three groups: known COVID-19 positive (n=8), unknown COVID-19 positive (n=15), and COVID-19 negative (n=18). All groups had similar pre-sample nucleocapsid (N) IgG levels as well as anti-spike IgG levels across WT, Delta, and Omicron SARS-CoV-2 strains. The known COVID-19 group had significantly higher post-sample N IgG (2.68 AU/ml) in comparison with the unknown positive (0.39 AU/ml) and negative (0.14 AU/ml) groups. Post sample levels of anti-spike IgG against WT, Delta, and Omicron were not significantly different between the known (66.4, 49.9, 21.1 AU/ml) and unknown (60.9, 41.9, 15.3 AU/ml) COVID-19 positive groups. Both groups were significantly increased across all strains in comparison to the COVID-19 negative group (25.1, 15.9, 5.4 AU/ml). *p<0.05 by two-way ANOVA.

Antibodies against spike protein increased from pre- to post-booster, as expected, since all participants received a booster dose immediately after baseline blood sampling. However, 120-day post-booster anti-spike IgG levels were significantly lower for the SARS-CoV-2 negative group in comparison with the known and unknown COVID-19+ groups (**Figure 1**). IgG levels did not differ between the known and unknown COVID-19+ groups. For all groups, anti-spike IgG levels were highest against WT followed by Delta and then Omicron.

To document the rate of antibody decline following the administration of the booster dose in the absence of infection, anti-spike IgG concentrations were analyzed in serially collected samples from participants in the COVID-19 negative group. Approximately 7 days after booster administration, anti-spike IgG levels increased against SARS-CoV-2 WT by 12.9-fold, Delta by 9.2-fold, and Omicron by 16.2-fold, compared to pre-booster levels (**Figure 2**). The concentration of WT anti-spike IgG was higher than IgG levels against Delta (p<0.05) and Omicron (p<0.0001) at 7 days post-booster administration and remained higher than both variant strains over the ∼120 days of sampling (**Figure 2**). At 120 days post-booster, median anti-spike IgG levels declined by 45% for antibodies against WT, 59% against Delta and 65% against Omicron compared to day 7 post-booster IgG levels.

**Figure 2.**
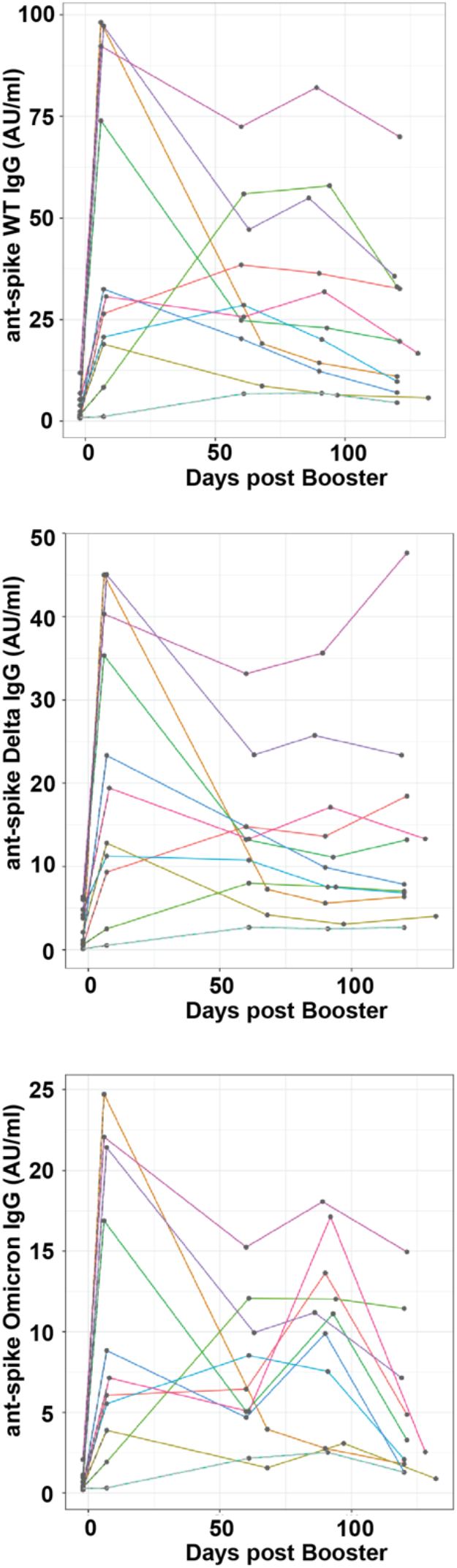
Rapid decline of anti-spike IgG across SARS-CoV-2 variants in uninfected persons. Matched, longitudinally collected DBS samples were evaluated for anti-spike IgG antibody levels against WT, Delta, and Omicron variants. Graphs illustrate individual level IgG responses with grey dots representing sample collection (n=11). Median antibody levels significantly increased 7 days post booster administration compared to pre booster levels with WT by 12.9-fold (pre 2.4, day 7 30.7 AU/ml), Delta by 9.2-fold (pre 2.1, day 7 19.4 AU/ml), and Omicron by 16.2-fold (pre 0.4, day 7 7.1 AU/ml); p < 0.002. Anti-spike IgG levels declined 120 days after booster with WT 45% (16.70 AU/ml), Delta 59% (7.9 AU/ml) and Omicron 65% (2.5 AU/ml) compared to day 7; with WT levels being significantly reduced by Friedman test p<0.006.

Due to the undetermined and variable timing of Omicron exposure among the known and unknown COVID+ groups it was not possible to report comparable rates of antibody decline for these groups. However, it is worth noting that at the conclusion of the four-month post-booster follow up period, the known and unknown COVID+ groups had anti-spike IgG concentrations that were 2.4 to 3.9 times higher than the SARS-CoV-2 negative group (**Figure 1**). Furthermore, anti-spike IgG concentrations were comparable for participants with unknown COVID-19 infection following the booster in comparison with participants with serologically-confirmed, self-reported COVID-19 infection after the booster.

## Discussion

In this prospective cohort study conducted during a window in which participants received booster vaccination prior to the Omicron wave, we document a high rate of Omicron infection after a booster, with both self-reported symptomatic cases and unknown infections based on anti-N seropositivity in both groups. Anti-spike antibodies recognizing WT, Delta, and Omicron variants all increased 9.2 to 16.2-fold after booster vaccination, but decreased rapidly over the subsequent four months. The key result is that both known and unknown COVID-19 infections during this period produced an additional, and comparable, boost in anti-spike antibody concentrations.

Recent estimates suggest that 57% of Americans were infected with Omicron due to its high transmissibility and potential for immune escape [3-5, 10]. In our cohort of recently boosted participants, one in five reported known infection based on a positive COVID-19 diagnostic test. Nearly twice as many (36.5%) had serological evidence of exposure in the absence of positive diagnostic testing, attesting to the high transmissibility of the Omicron variant and the utility of using anti-N antibody testing to more fully assess prevalence. Importantly, anti-spike antibody levels were comparable in the known and unknown COVID-19+ groups at the end of the follow up period and higher than those who remained SARS-CoV-2 uninfected, indicating that symptomatic infection is not necessary to generate a robust antibody response in vaccinated and boosted persons.

While COVID-19 vaccines are very effective at preventing severe disease, our serological results reveal that infections following a booster are more common than what is ascertained from diagnostic testing. Neutralizing antibody levels peak soon after vaccination and decline over time, increasing the risk for asymptomatic or relatively mild infections beginning approximately 4 months following the second dose of mRNA vaccines [11]. Our study demonstrates the effectiveness of boosters in increasing antibodies, but also shows rapid rates of decline, particularly for anti-spike IgG antibodies against the Omicron variant. To the best of our knowledge, our study is the first to report that post-booster infections increase antibody levels above and beyond what is seen with a booster dose in known as well as unknown infections. However, the anti-spike IgG response against Omicron is significantly smaller than responses against other variants, and the rate of decline is faster, which may indicate a lower level of protection against variants of this lineage.

Limitations of our study include the small sample size, and the absence of measures of cellular immunity to complement our assessment of antibody concentrations. In addition, our cohort includes fully vaccinated and boosted individuals, which precludes us from evaluating the likelihood of infection and magnitude of antibody response among the unvaccinated and those vaccinated with the initial doses but not boosted.

## Data Availability

Written requests for data should be made by qualified researchers trained in human subject confidentiality protocols to the corresponding author(s).

## FIGURES and LEGENDS

**SUPPLEMENTAL TABLE 1.** Cohort demographics.

|  | Known COVID-19+ |  | Unknown COVID-19+ |  | Negative COVID-19 |  | Total |
| --- | --- | --- | --- | --- | --- | --- | --- |
|  | n | % | n | % | n | % | n |
| **Total participants** | 8 |  | 15 |  | 18 |  | 41 |
| **Birth Sex** |  |  |  |  |  |  |  |
| Female | 5 | 62.5 | 7 | 46.7 | 9 | 50.0 | 21 |
| Male | 3 | 37.5 | 8 | 53.3 | 9 | 50.0 | 20 |
| **Age (median, yrs)** | 45 |  | 39 |  | 39 |  | 41 |
| range (yrs) | 25 to 66 |  | 23 to 66 |  | 22 to 95 |  | 22 to 95 |
| **Vaccine Manufacturer** |  |  |  |  |  |  |  |
| Pfizer | 3 | 37.5 | 10 | 66.7 | 12 | 66.7 | 25 |
| Moderna | 5 | 62.5 | 5 | 33.3 | 6 | 33.3 | 16 |
| **Days between booster and post sample (median)** | 122 |  | 120 |  | 120 |  | 120 |
| **Known Exposed Household** | 4 | 50 | 6 | 40.0 | 2 | 11.1 | 12 |

